# Prevalence of incidental germline variants detected via tumor-only mesothelioma genomic profiling

**DOI:** 10.1101/2022.12.06.22282680

**Authors:** Owen D. Mitchell, Katie Gilliam, Daniela del Gaudio, Kelsey E. McNeely, Shaili Smith, Maria Acevedo, Meghana Gaduraju, Rachel Hodge, Aubrianna S.S. Ramsland, Jeremy Segal, Soma Das, Darren S. Bryan, Sanjukta Tawde, Shelly Galasinski, Peng Wang, Melissa Y. Tjota, Aliya N. Husain, Samuel Armato, Jessica Donington, Mark K. Ferguson, Kiran Turaga, Jane E. Churpek, Hedy L. Kindler, Michael W. Drazer

**Affiliations:** Section of Hematology/Oncology, Department of Medicine, The University of Chicago, Chicago, IL, USA; Department of Human Genetics, The University of Chicago, Chicago, IL, USA; Department of Pathology, The University of Chicago, Chicago, IL, USA; Department of Radiology, The University of Chicago, Chicago, IL, USA; Department of Surgery, The University of Chicago, Chicago, IL, USA; Division of Hematology, Medical Oncology, and Palliative Care, Department of Medicine, University of Wisconsin, Madison, WI, USA

## Abstract

**Importance:** Patients with mesothelioma often have next generation sequencing (NGS) of their tumor. Tumor-only NGS may incidentally identify germline pathogenic or likely pathogenic (P/LP) variants despite not being designed for this purpose. It is unknown how frequently patients with mesothelioma have germline P/LP variants incidentally detected via tumor-only NGS.

**Objective:** To determine the prevalence of incidental germline P/LP variants detected via tumor-only NGS of mesothelioma.

**Design:** A series of 161 unrelated patients with mesothelioma had tumor-only NGS and germline NGS. These assays were compared to determine which P/LP variants identified via tumor-only NGS were of germline origin.

**Setting:** This was an observational study from a high-volume mesothelioma program. All patients enrolled on Institutional Review Board-approved protocols.

**Participants:** 161 unrelated patients with pleural, peritoneal, or bicavitary mesothelioma.

**Intervention(s) (for clinical trials) or Exposure(s) (for observational studies):** No therapeutic interventions were used.

**Main Outcome(s) and Measure(s):** The proportion of patients with mesothelioma who had P/LP germline variants incidentally detected via tumor-only NGS.

**Results:** Most (78%) patients had potentially incidental P/LP germline variants. The positive predictive value of a potentially incidental germline P/LP variant on tumor-only NGS was 20%. Overall, 16% of patients carried a P/LP germline variant. Germline P/LP variants were identified in *ATM, ATR, BAP1, CHEK2, DDX41, FANCM, HAX1, MRE11A, MSH6, MUTYH, NF1, SAMD9L*, and *TMEM127*.

**Conclusions and Relevance:** Most (78%) patients with mesothelioma had potentially incidental germline P/LP variants on tumor NGS, but the positive predictive value of these was modest (20%). Of all patients, 16% had confirmed germline P/LP variants. Given the implications of a hereditary cancer syndrome diagnosis for preventive care and familial counseling, clinical approaches for addressing incidental P/LP germline variants in tumor-only NGS are needed.Tumor-only sequencing should not replace dedicated germline testing. Universal germline testing is likely needed for patients with mesothelioma.

**Key Points:** *Question:* What proportion of patients with mesothelioma have pathogenic or likely pathogenic germline variants incidentally identified by tumor-only genomic profiling?

*Findings:* In this cohort study of 161 patients with mesothelioma, 78% of patients had potential germline variants that warranted further evaluation. Overall, 16% of patients had pathogenic or likely pathogenic germline variants initially identified via tumor-only genomic profiling.

*Meaning:* Tumor genomic profiling of mesothelioma frequently (78% of patients) identifies potential germline pathogenic/likely pathogenic variants that warrant dedicated germline evaluation. The high prevalence of germline variants (16%) in our series suggests universal genetic testing may be warranted for patients with mesothelioma.

## Introduction

Mesothelioma is an aggressive cancer that principally affects the pleural and/or peritoneal cavities.^1^ The prognosis of mesothelioma is poor, with a median survival of only 18.4 months.^2^ Anatomic tumor location does influence prognosis, as individuals with peritoneal mesothelioma have longer survival compared to patients with pleural mesothelioma.^3^

Asbestos exposure is the major known risk factor for mesothelioma, but 12% of patients also carry germline pathogenic or likely pathogenic variants (P/LP variants) that further modify an individual’s lifetime risk of solid tumor development.^3-7^ Individuals with peritoneal mesothelioma are particularly likely to carry germline P/LP variants (25% of cases).^4^ Universal germline genetic testing, which is increasingly a standard of care in other cancers, such as epithelial ovarian cancer and exocrine pancreatic cancer, is not yet routinely employed in mesothelioma care.^8^ Clinical guidelines for mesothelioma recommend discussing the risks and benefits of germline genetic testing with patients who have personal or family histories that are suggestive of a hereditary cancer syndrome, particularly if these malignancies are associated with germline *BAP1* P/LP variants.^9^ Experiences with other tumor types have shown that universal genetic testing, as opposed to germline testing triggered by high risk personal and/or family histories, increases the diagnosis of hereditary cancer syndromes.^10^

Germline *BAP1* P/LP variants are the most frequent and well-studied alterations in patients with mesothelioma who have increased cancer risk. Patients with mesothelioma and germline P/LP variants, especially in *BAP1*, have improved survival relative to patients without germline P/LP variants.^3, 11^ Recognition of patients with germline *BAP1* P/LP variants may also help guide treatment decision making, particularly in regard to the use of platinum-based therapies.^3, 4, 9^

While germline genetic testing remains variably implemented for patients with mesothelioma, tumor-only next-generation sequencing (NGS) is increasingly being used in clinical practice, especially in academic settings. This utilization is driven, in part, by the number of novel therapies that are being developed to target dysregulated genes and pathways, such as the EZH2 protein and the Hippo pathway.^12, 13^ Tumor-only NGS may also help identify clinical trials for which patients may be eligible. Some of the most common somatically mutated genes in mesothelioma include *BAP1, CDKN2A, DDX3X, NF2*, and *TP53*.^14, 15^ These same genes, when mutated in the germline, may predispose to a variety of solid tumors (*BAP1, CDKN2A, NF2, TP53)* as well as developmental delays and/or disabilities (*DDX3X*).^9, 16-20^ Recognizing the germline origin of these variants has important implications for the counseling and care of both the index patient and their family members.^9^ Some P/LP variants detected via tumor-only NGS may actually be of germline origin, but the correlation between tumor-only NGS and germline sequencing has not previously been performed for patients with mesothelioma.

Here, we analyzed a series of patients with mesothelioma who underwent both tumor-only NGS and germline sequencing. We aimed to determine the prevalence of incidental germline findings that are detected via tumor-only sequencing.

## Results

### Study Population

168 unrelated patients with mesothelioma who had tumor-only NGS were included in this study. Of these patients, 161 (96%) had sufficient germline tissue available for sequencing (**Figure 1**). Approximately two-thirds of the patients were male and one-third were female. Most (156 of 161; 97%) patients self-identified as non-Hispanic white. Approximately 70% of all patients had pleural mesothelioma, 29% had peritoneal mesothelioma, and 1% had bicavitary disease or involvement of the tunica vaginalis. The majority (86%) of patients had an epithelioid subtype of mesothelioma, with biphasic (9%), sarcomatoid (5%) and benign multicystic (0.01%) histologies representing a minority of patients. Most patients (76%) did not have a personal history of second cancers. Among the 39 patients (24%) with second malignancies, 12 had a history of skin cancer, 8 had prostate cancer, 7 had lymphoma, and 4 had thyroid cancer. 71% of all patients had a first-degree family member with a malignancy (**Table 1**).

**Table 1.** Characteristics of mesothelioma patients and tumors

| **Table 1. Characteristics of mesothelioma patients and tumors** |  |  |  |  |  |
| --- | --- | --- | --- | --- | --- |
| Characteristic | Patients with P or LP<br>Germline Variants |  | Patients without P or LP<br>Germline Variants |  | p-value |
|  | n | [%] | n | [%] |  |
| Gender |  |  |  |  |  |
| Male | 16 | 64% | 89 | 64% | 1 |
| Female | 9 | 36% | 47 | 35% |  |
| Age at Diagnosis (y) |  |  |  |  |  |
| <20 | 0 | 0% | 0 | 0% | 0.9196 |
| 20-29 | 0 | 0% | 2 | 1% |  |
| 30-39 | 0 | 0% | 1 | 1% |  |
| 40-49 | 1 | 4% | 7 | 5% |  |
| 50-59 | 6 | 24% | 31 | 22% |  |
| 60-69 | 9 | 36% | 51 | 38% |  |
| 70-79 | 6 | 24% | 36 | 26% |  |
| >80 | 3 | 12% | 8 | 6% |  |
| Ethnicity |  |  |  |  |  |
| White, non-Hispanic | 25 | 100% | 131 | 96% | 1 |
| White, Hispanic | 0 | 0% | 1 | 1% |  |
| Black | 0 | 0% | 1 | 1% |  |
| Asian/South Asian | 0 | 0% | 3 | 2% |  |
| Asbestos Exposure* |  |  |  |  |  |
| Definite | 10 | 40% | 52 | 38% | 0.7296 |
| Probable | 4 | 16% | 32 | 24% |  |
| Possible | 8 | 32% | 41 | 30% |  |
| No Known Exposure | 3 | 12% | 10 | 7% |  |
| No Data | 0 | 0% | 1 | 1% |  |
| Mesothelioma Site |  |  |  |  |  |
| Pleura | 15 | 60% | 94 | 69% | 0.6119 |
| Peritoneum | 9 | 36% | 36 | 26% |  |
| Bicavitary | 1 | 4% | 4 | 3% |  |
| Tunica vaginalis | 0 | 0% | 2 | 1% |  |
| Pericardium | 0 | 0% | 0 | 0% |  |
| Tumor Histology |  |  |  |  |  |
| Epithelial | 23 | 92% | 115 | 85% | 0.6854 |
| Sarcomatoid | 0 | 0% | 8 | 6% |  |
| Biphasic | 2 | 8% | 12 | 9% |  |
| Benign Multicystic | 0 | 0% | 1 | 1% |  |
| Personal History of Other Cancer |  |  |  |  |  |
| Yes | 12 | 48% | 27 | 18% | **0.0047** |
| No | 13 | 52% | 109 | 81% |  |
| Other Cancers** |  |  |  |  |  |
| Colon | 0 | 0% | 2 | 6% |  |
| Lymphoma | 3 | 19% | 4 | 13% |  |
| Prostate | 2 | 13% | 6 | 19% |  |
| Renal | 2 | 13% | 0 | 0% |  |
| Skin, Not Otherwise Specified | 3 | 19% | 9 | 29% |  |
| Thyroid | 1 | 6% | 3 | 10% |  |
| Other | 5 | 31% | 7 | 23% |  |
| Cancer in 1st degree relative |  |  |  |  |  |
| Yes | 23 | 92% | 91 | 68% | **0.0148** |
| No | 2 | 8% | 45 | 33% |  |

**Figure 1:**
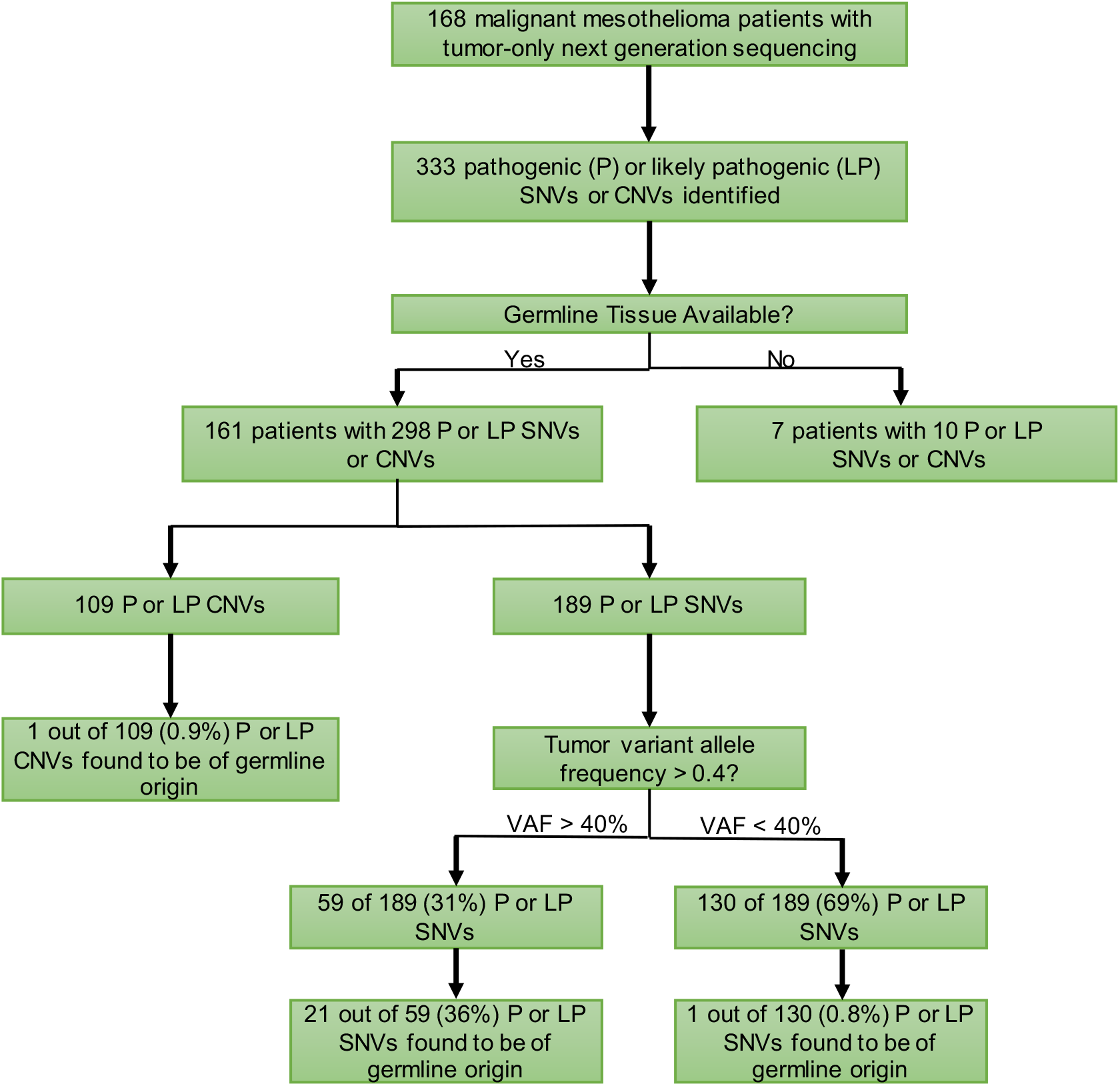
CONSORT diagram of patients in mesothelioma sequencing series.

### Somatic and Germline Pathogenic/Likely Pathogenic Variants

Most (126 of 161; 78%) patients with tumor-only NGS had a P/LP variant detected in at least one gene associated with a hereditary cancer syndrome, while only 25 of the 161 patients (16%) were ultimately found to carry a P/LP germline variant in at least one of 84 genes sequenced via germline NGS (**Figure 1**). A total of 28 germline P/LP variants were identified, and three of the 25 patients carried P/LP germline variants in two different genes. Most germline variants were in genes involved in DNA repair: *BAP1* (n=8, 29% of patients with a germline P/LP variant), *CHEK2* (n=6, 21%) and *ATM* (n=3, 11%). Other genes with a germline P/LP variant identified in at least one patient included: *ATR, DDX41, FANCM, HAX1, MRE11A, MSH6, MUTYH, NF1, SAMD9L*, and *TMEM127* (**Table 2, Supp. Fig 1**. Each patient with multiple P/LP germline variants carried a P/LP *BAP1* variant in addition to germline P/LP variants in additional genes: *TMEM127* (Patient 8), *HAX1* (Patient 9), and *SAMD9L* (Patient 10). The positive predictive value (PPV) of a P/LP variant identified on tumor-NGS was 20% (n=25/126).

**Table 2.**
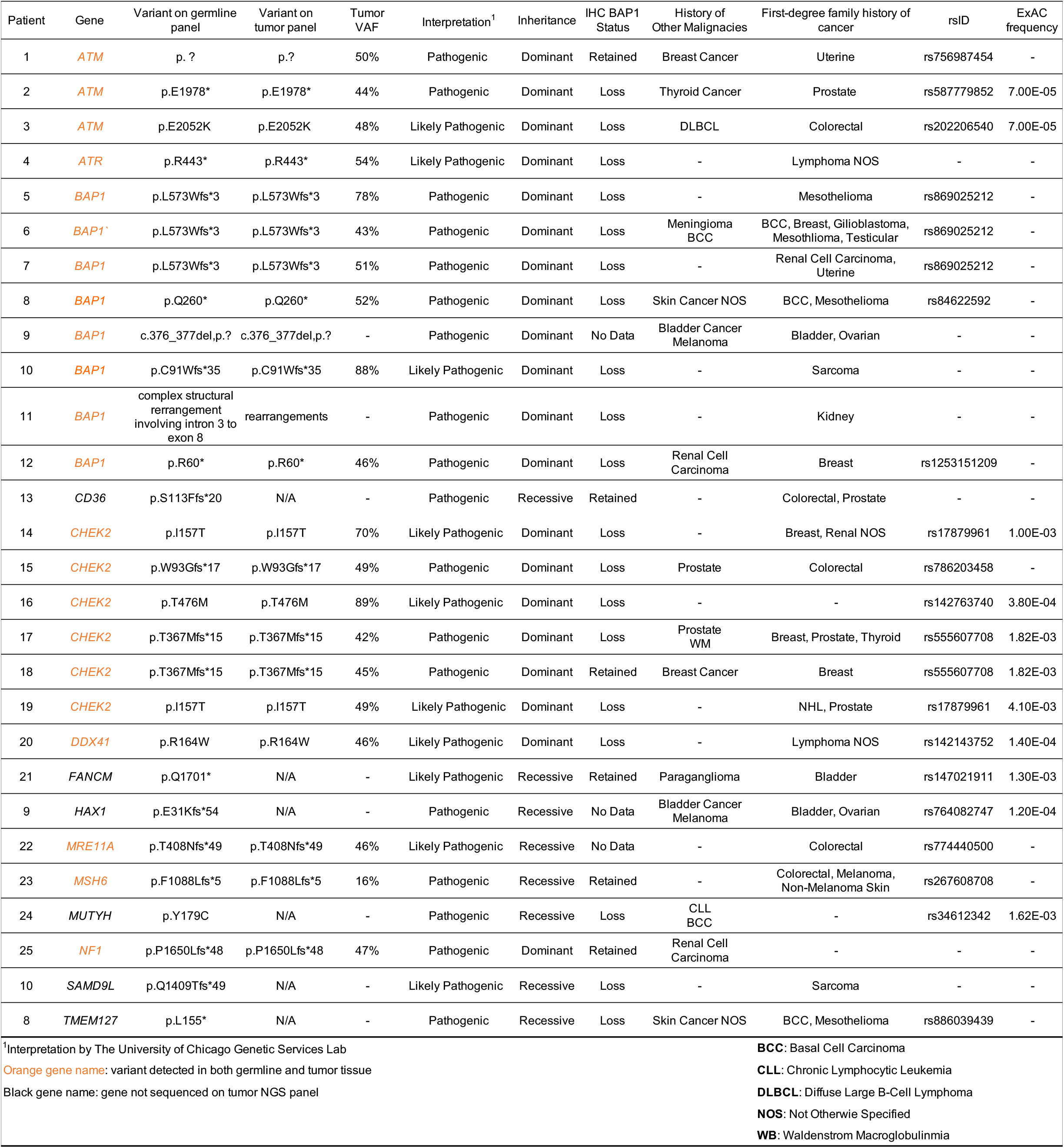
Patients in cohort with a germline pathogenic / likely pathogenic germline variant.

| Patient | Gene | Variant on germline panel | Variant on tumor panel | Tumor VAF | Interpretation<sup>1</sup> | Inheritance | IHC BAP1 Status | History of Other Malignancies | First-degree family history of cancer | rsID | ExAC frequency |
| --- | --- | --- | --- | --- | --- | --- | --- | --- | --- | --- | --- |
| 1 | *ATM* | p.? | p.? | 50% | Pathogenic | Dominant | Retained | Breast Cancer | Uterine | rs756987454 | - |
| 2 | *ATM* | p.E1978* | p.E1978* | 44% | Pathogenic | Dominant | Loss | Thyroid Cancer | Prostate | rs587779852 | 7.00E-05 |
| 3 | *ATM* | p.E2052K | p.E2052K | 48% | Likely Pathogenic | Dominant | Loss | DLBCL | Colorectal | rs202206540 | 7.00E-05 |
| 4 | *ATR* | p.R443* | p.R443* | 54% | Likely Pathogenic | Dominant | Loss | - | Lymphoma NOS | - | - |
| 5 | *BAP1* | p.L573Wfs*3 | p.L573Wfs*3 | 78% | Pathogenic | Dominant | Loss | - | Mesothelioma | rs869025212 | - |
| 6 | *BAP1* | p.L573Wfs*3 | p.L573Wfs*3 | 43% | Pathogenic | Dominant | Loss | Meningioma BCC | BCC, Breast, Glioblastoma, Mesothelioma, Testicular | rs869025212 | - |
| 7 | *BAP1* | p.L573Wfs*3 | p.L573Wfs*3 | 51% | Pathogenic | Dominant | Loss | - | Renal Cell Carcinoma, Uterine | rs869025212 | - |
| 8 | *BAP1* | p.Q260* | p.Q260* | 52% | Pathogenic | Dominant | Loss | Skin Cancer NOS | BCC, Mesothelioma | rs84622592 | - |
| 9 | *BAP1* | c.376_377del,p.? | c.376_377del,p.? | - | Pathogenic | Dominant | No Data | Bladder Cancer Melanoma | Bladder, Ovarian | - | - |
| 10 | *BAP1* | p.C91Wfs*35 | p.C91Wfs*35 | 88% | Likely Pathogenic | Dominant | Loss | - | Sarcoma | - | - |
| 11 | *BAP1* | complex structural rearrangement involving intron 3 to exon 8 | rearrangements | - | Pathogenic | Dominant | Loss | - | Kidney | - | - |
| 12 | *BAP1* | p.R60* | p.R60* | 46% | Pathogenic | Dominant | Loss | Renal Cell Carcinoma | Breast | rs1253151209 | - |
| 13 | *CD36* | p.S113Ffs*20 | N/A | - | Pathogenic | Recessive | Retained | - | Colorectal, Prostate | - | - |
| 14 | *CHEK2* | p.I157T | p.I157T | 70% | Likely Pathogenic | Dominant | Loss | - | Breast, Renal NOS | rs17879961 | 1.00E-03 |
| 15 | *CHEK2* | p.W93Gfs*17 | p.W93Gfs*17 | 49% | Pathogenic | Dominant | Loss | Prostate | Colorectal | rs786203458 | - |
| 16 | *CHEK2* | p.T476M | p.T476M | 89% | Likely Pathogenic | Dominant | Loss | - | - | rs142763740 | 3.80E-04 |
| 17 | *CHEK2* | p.T367Mfs*15 | p.T367Mfs*15 | 42% | Pathogenic | Dominant | Loss | Prostate WM | Breast, Prostate, Thyroid | rs555607708 | 1.82E-03 |
| 18 | *CHEK2* | p.T367Mfs*15 | p.T367Mfs*15 | 45% | Pathogenic | Dominant | Retained | Breast Cancer | Breast | rs555607708 | 1.82E-03 |
| 19 | *CHEK2* | p.I157T | p.I157T | 49% | Likely Pathogenic | Dominant | Loss | - | NHL, Prostate | rs17879961 | 4.10E-03 |
| 20 | *DDX41* | p.R164W | p.R164W | 46% | Likely Pathogenic | Dominant | Loss | - | Lymphoma NOS | rs142143752 | 1.40E-04 |
| 21 | *FANCM* | p.Q1701* | N/A | - | Likely Pathogenic | Recessive | Retained | Paraganglioma | Bladder | rs147021911 | 1.30E-03 |
| 9 | *HAX1* | p.E31Kfs*54 | N/A | - | Pathogenic | Recessive | No Data | Bladder Cancer Melanoma | Bladder, Ovarian | rs764082747 | 1.20E-04 |
| 22 | *MRE11A* | p.T408Nfs*49 | p.T408Nfs*49 | 46% | Likely Pathogenic | Recessive | No Data | - | Colorectal | rs774440500 | - |
| 23 | *MSH6* | p.F1088Lfs*5 | p.F1088Lfs*5 | 16% | Pathogenic | Recessive | Retained | - | Colorectal, Melanoma, Non-Melanoma Skin | rs267608708 | - |
| 24 | *MUTYH* | p.Y179C | N/A | - | Pathogenic | Recessive | Loss | CLL BCC | - | rs34612342 | 1.62E-03 |
| 25 | *NF1* | p.P1650Lfs*48 | p.P1650Lfs*48 | 47% | Pathogenic | Dominant | Retained | Renal Cell Carcinoma | - | - | - |
| 10 | *SAMD9L* | p.Q1409Tfs*49 | N/A | - | Likely Pathogenic | Recessive | Loss | - | Sarcoma | - | - |
| 8 | *TMEM127* | p.L155* | N/A | - | Pathogenic | Recessive | Loss | Skin Cancer NOS | BCC, Mesothelioma | rs886039439 | - |

<sup>1</sup>Interpretation by The University of Chicago Genetic Services Lab
Orange gene name: variant detected in both germline and tumor tissue
Black gene name: gene not sequenced on tumor NGS panel
**BCC:** Basal Cell Carcinoma
**CLL:** Chronic Lymphocytic Leukemia
**DLBCL:** Diffuse Large B-Cell Lymphoma
**NOS:** Not Otherwise Specified
**WB:** Waldenstrom Macroglobulinemia

### Characteristics of Germline Pathogenic/Likely Pathogenic Variant Carriers

Most patients (n=15, 60%) with P/LP germline variants had pleural mesothelioma, 36% (n=9) had peritoneal mesothelioma, and 4% (n=1) had bicavitary disease. Asbestos exposure was not significantly different between germline P/LP variant carriers and patients without germline variants, with “Definite” being the most reported asbestos exposure in both groups (40% *vs* 38%, *p*=0.85). Germline variant carriers were more likely (*p*=0.001) to have had a second cancer (n=12 of 25, 48%) as compared to patients without germline variants (n=27 of 136, 18%). Most (11 of 12, 92%) patients with germline P/LP germline variants who had multiple cancers were diagnosed with mesothelioma concurrently or after the diagnosis of a second malignancy (**Supp Fig 2**). The average time between a first diagnosis of cancer and a mesothelioma diagnosis in patients with P/LP germline variants was 9.8 years (**Supp. Fig 2**). This prolonged survival period indicates that these patients may potentially benefit from cancer screening if a hereditary cancer syndrome is recognized at the time of their first cancer diagnosis. Germline P/LP variant carriers were more likely (*p*=0.001) to have at least one first-degree family member with a cancer diagnosis (n=23, 92%) than patients without a germline P/LP variant (n=91, 68%, **Table 1**).

### Pathologic Characteristics of Germline Pathogenic/Likely Pathogenic Variant Carriers

Overall, 23 germline P/LP variant carriers had mesothelioma with epithelioid histology and two germline P/LP variant carriers had biphasic histology. These proportions were similar to patients without a germline variant (85% epithelioid and 9% biphasic or sarcomatoid, respectively, *p*=0.35 and *p*=0.37, **Table 1**). Immunohistochemistry showed a majority of tumors in germline P/LP variant carriers (n=17, 68%) lost BAP1 expression, which was similar (*p*=0.28) to patients without germline P/LP variants (n=69, 51%). Programmed death-ligand one (PD-L1) staining was positive in 56% (n=14) of germline P/LP variant carrier tumors, which was similar (*p*=0.38) to patients without germline P/LP variants (n = 61, 45%). Of the 14 PD-L1 positive tumors in the P/LP germline variant carriers, half had a tumor proportion score (TPS) of less than 5% (n=7) and staining intensity was classified as either weak (n=5) or weak-to-moderate. Interestingly, most patients with evaluable *CHEK2* germline P/LP variant carriers (n=5 of 6; 83%) had tumors with BAP1 loss and positive PD-L1 staining (**Supp Fig 3**). All evaluable tumors from *BAP1* germline P/LP variant carriers (n=7) had BAP1 loss, but PD-L1 expression was positive in four and negative in three tumor samples.

### Co-Mutational Plot of Somatic and Germline Pathogenic/Likely Pathogenic Variants

All known germline P/LP variants in the patient cohort were incidentally detected via tumor-only NGS panels that sequenced the same genes (**Supp Fig 4**). A small number of patients had P/LP germline variants detected via dedicated germline sequencing only, as the genes of interest (*CD36, FANCM, HAX1, MUTYH, SAMD9L, TMEM127*) were not sequenced by our institution’s tumor-only NGS panel. The tumor variant allele frequency (VAF) for incidental germline P/LP variants was greater than 40% in 19 of the 20 variants for which VAF was available (**Table 2**). Only one germline variant, a frameshift variant in *MSH6*, had a tumor VAF less than 40% (**Table 2**). The most frequent somatic mutations in patients with germline P/LP variants were located in *DDX3X* (n=4), *NF2* (n=4), and *TP53* (n=3, **Figure 2**). Three of eight patients (38%) with germline P/LP variants in *BAP1* had second somatic hits in *BAP1*. The VAF of these second hits in *BAP1* ranged from 18% to 25%.

**Figure 2.**
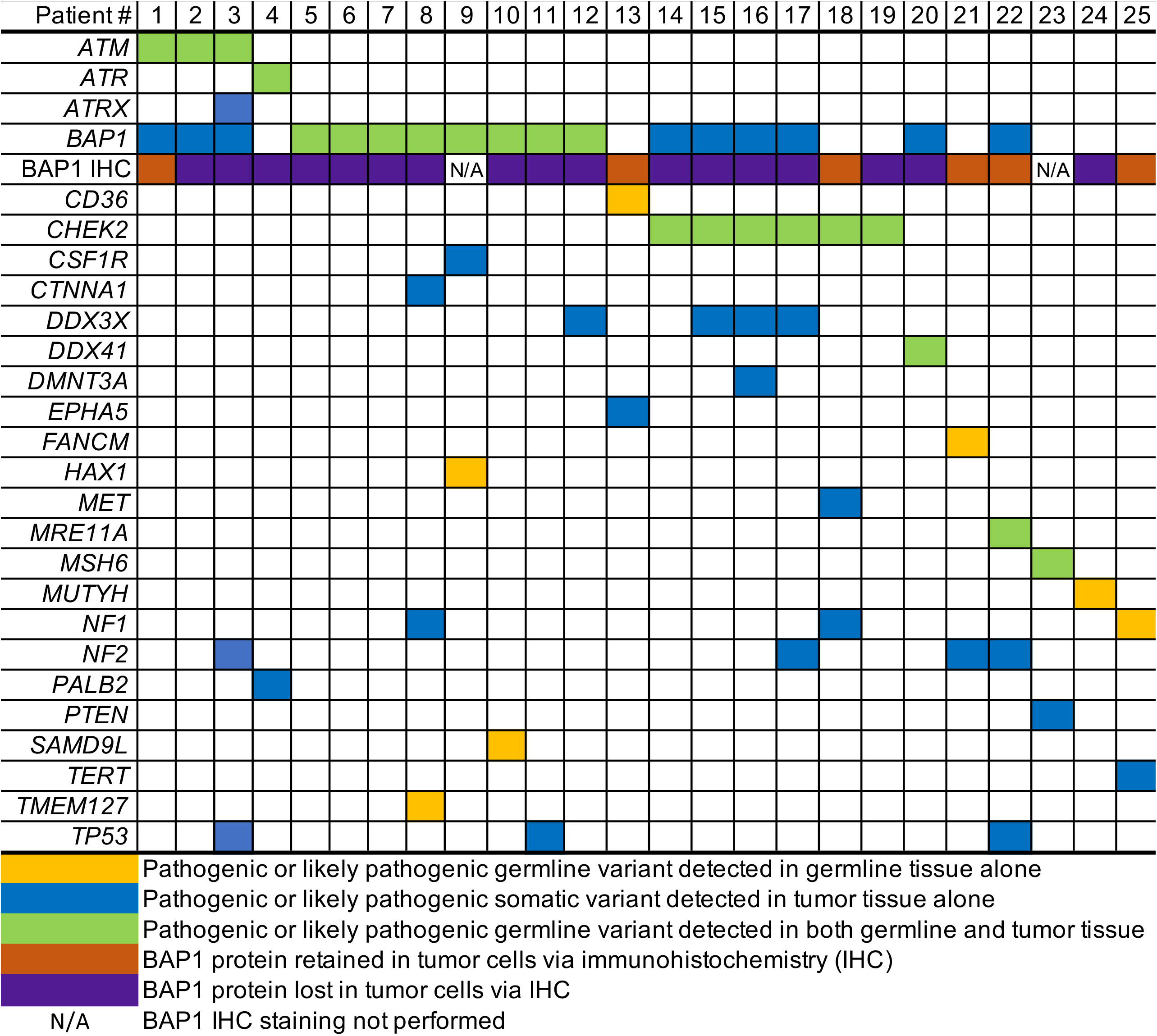
Pathogenic or Likely Pathogenic Variants in Patients with Germline Mutations

## Discussion

The role of P/LP germline variants in modifying cancer risk, as well as tumor prognosis and treatment strategies, in unselected cohorts of oncology patients is increasingly recognized. Large sequencing cohorts of otherwise unselected oncology patients with a wide variety of tumor types that lack formal hereditary testing guidelines (bladder, brain, cholangiocarcinoma, esophagus, head/neck, lung, and others) have shown that approximately 7% of patients with these tumor types carry germline P/LP variants.^21^ The germline diagnostic yield for germline P/LP variants is particularly high in other tumors, such as pancreatic and ovarian cancers, for which established hereditary testing guidelines exist.^8^ Mesothelioma is now recognized as a cancer that is similarly enriched for hereditary cancer syndromes, with 12-16% of patients with mesothelioma carrying P/LP germline variants.^3-7^

Similarly, the rapid uptake of tumor-based NGS has shown that many patients with otherwise unremarkable personal and family histories will have germline P/LP variants incidentally detected via tumor-only NGS, despite these assays not being dedicated germline tests.^22-24^ Studies of incidental germline findings have occurred in multiple tumor types, but no study to date has examined the rate of incidental findings on tumor-only NGS for patients with mesothelioma.^23-29^

Here, we demonstrate that 78% of patients with mesothelioma have P/LP variants detected via tumor-only sequencing that meet NCCN criteria for dedicated germline testing. The PPV of a potentially incidental germline P/LP variant on tumor-only testing, however, was modest (20%), suggesting that many patients who undergo tumor-only NGS will have variants of potential germline origin that warrant further evaluation. Overall, 16% (n=25/161) of individuals in our unselected series of patients with mesothelioma carried P/LP germline variants that were first detected incidentally via tumor-only sequencing. The overall yield of incidentally detected germline P/LP variants in our series (16%) is similar to the germline yield (12%) found using a universal germline sequencing approach in an unselected cohort of patients with mesothelioma from our institution.^4, 30^ Some patients were identified in both cohorts. Overall, these findings suggest that the germline diagnostic yield for patients with mesothelioma is similar to other tumor types in which universal germline genetic testing is recommended by NCCN guidelines. Given this, it is reasonable that future clinical guidelines advocate that universal germline sequencing be offered to patients with mesothelioma.

Importantly, 4.0% (n=6 of 161) of patients in our cohort had germline P/LP variants detected via germline sequencing only, but not via tumor-only NGS. This phenomenon occurred as our institution’s tumor-only NGS panel did not sequence the germline gene of interest. This finding reinforces the point that tumor-only NGS should not replace dedicated germline testing, as these patients would be placed at risk for “false negative” testing in these situations.

Standards of care in regard to germline genetic testing in patients with mesothelioma, however, remain unclear. Multiple groups have proposed varied approaches to germline testing in patients with cancer. One proposed approach would be to perform universal germline testing of all cancer patients, particularly for patient populations whose tumors exceeding a pre-defined threshold for cancer risk (i.e., 10% of all patients will be expected to carry germline P/LP variants).^10, 31, 32^ Another recently proposed approach would be to first consider germline testing based on the patient’s personal/family history, in patients with microsatellite unstable tumors, or in patients with a potential germline P/LP variant detected via tumor-only NGS.^33^ In our series, the majority of patients with mesothelioma meet one of these aforementioned criteria.

We found very few clinical-level factors, aside from family history and the presence of a potentially incidental germline variant on tumor-only NGS, predicted for the presence of a germline P/LP variant. We did find an increased likelihood that patients whose family members had been diagnosed with cancer were carriers of germline P/LP variants (*p* < 0.001). However, we also identified germline variants in a number of patients who did not have a strong family history of cancer (**Table 1**), suggesting that family history alone was not a reliable predictor for the presence of a germline P/LP variant. Mesothelioma is a unique and rare cancer for which asbestos exposure is a major risk factor.^34, 35^ In our series, rates of asbestos exposure were statistically equivalent between patients with and without germline P/LP variants. Furthermore, mesothelioma is a highly microsatellite stable malignancy, which limits the utility of using microsatellite instability as a guide for germline testing.^36^ In addition, the genes that were most frequently somatically mutated in mesothelioma tumor tissue are frequently involved in hereditary cancer syndromes, as evidenced by the modest positive predictive value of potentially incidental findings in our study (20%). Finally, tumor VAF has been proposed as a potential surrogate for germline variants.^33^ Even though our analysis found that the majority (21 of 22; 95%) of germline variants on tumor-only NGS had a VAF greater than 40%, the importance of avoiding “false negative” germline evaluations suggests that VAF alone should not be used as a surrogate for dedicated germline testing (**Figure 1**).

Based on the high prevalence of germline P/LP variants in patients with mesothelioma (12-16%), the high prevalence of variants on tumor-only NGS that warrant dedicated germline evaluation (78%), the inability to use tumor-based NGS as a surrogate, the clinical importance of identifying hereditary cancer syndromes, and the decreased cost of NGS, we are likely approaching an inflection point in which universal germline testing of patients with mesothelioma warrants consideration in clinical care guidelines.

## Methods

### Study Population

Patients with mesothelioma who presented to the University of Chicago Medicine (UCM) mesothelioma clinic were prospectively consented from April 2016 to October 2021. Peripheral blood, saliva and tumor specimens were collected. A personal and family history of cancer was obtained by trained interviewers using a standardized survey. Asbestos exposure was self-reported by patients, citing primary exposures (occupational, environmental) or secondary exposures (living with persons exposed to asbestos). All reported exposures were reviewed by researchers and then classified as definite, probable, possible, or no known exposure. Other demographic and clinical information was abstracted from the medical record. The UCM Institutional Review Board approved this study.

### Germline Mutation Detection & Interpretation

DNA was extracted from either saliva or peripheral blood in both Clinical Laboratory Improvement Amendments (CLIA) – certified and non-CLIA-certified laboratories. Extracted DNA was assayed on a research basis using a CLIA-certified targeted gene panel that was designed by the University of Chicago Genetic Services Laboratory to capture and sequence the coding and flanking intronic regions of 84 cancer susceptibility genes. All variants were analyzed by two independent reviewers and interpreted according to the American College of Medical Genetics and Genomics and Association for Molecular Pathology consensus guidelines.^37^ P/LP variants, including nonsense, frameshift, splice site, missense variants, and large-scale genomic rearrangements with known moderate-to-high penetrance cancer susceptibility were reported, in addition to variants of unknown significance (VUS). All germline P/LP variants were validated by Sanger sequencing, correlated with clinical and family history, and segregated in family members when possible.

### Somatic Mutation Detection & Interpretation

DNA was extracted from fresh-frozen, paraffin embedded tumor tissue blocks. Somatic mutations were identified using version six of the UCM OncoPlus next-generation sequencing panel (n = 1212 genes, clinically reported genes included in **Supp Fig 1**).^38^

### Immunohistochemistry

BAP1 and PD-L1 staining was conducted in a CLIA-certified laboratory and interpretation was performed by the attending pathologist and reviewed with a pathology resident or fellow. The Santacruz, C4 monoclonal antibody was used for BAP1 staining and the Abcam 28.8 monoclonal antibody was used to evaluate PD-L1 staining. Percentage of cells positive for PD-L1 was calculated via the Tumor Percentage Score (TPS).

### Statistical Analyses

Fisher’s exact test was used to determine an association between patients with mesothelioma, with and without germline mutations for patient and tumor characteristics. A *p-*value < 0.05 was considered to be statistically significant.

## Data Availability

All data produced in the present work are contained in the manuscript

## Acknowledgments

The authors thank the patients who participated in this research and donated biological specimens to the University of Chicago.

