## Supplemental Figure 1 for "Prevalence of incidental germline variants detected via tumor-only mesothelioma genomic profiling"

**Supplementary Table 1. Genes sequenced on the germline and somatic panels**

| GENES ON MESOTHELIOMA GERMLINE PANEL ONLY | | GENES ON TUMOR PANEL ONLY | | | | GENES ON BOTH PANELS | |
|---|---|---|---|---|---|---|---|
| *ANKR26* | NM_014915.2 | *ABL1* | NM_005157.6 | *HIST1H3C* | NM_003531.3 | *APC* | NM_000038.5 |
| *BMPR1A* | NM_004329.2 | *AKT1* | NM_001382430.1 | *HNF1A* | NM_000545.8 | *ATM* | NM_000051.3 |
| *BRIP1* | NM_032043.2 | *ALK* | NM_004304.5 | *HRAS* | NM_005343.4 | *ATR* | NM_001184.4 |
| *CD36* | NM_000072.3 | *ARID1A* | NM_006015.6 | *IDH1* | NM_005896.4 | *BAP1* | NM_004656.3 |
| *EPCAM* | NM_002354.2 | *ARID2* | NM_152641.4 | *IDH2* | NM_002168.4 | *BARD1* | NM_000465.4 |
| *ERCC4* | NM_005236.2 | *ASXL1* | NM_015338.6 | *ITPKB* | NM_002221.4 | *BLM* | NM_000057.3 |
| *FANCB* | NM_001018113.2 | *AXL* | NM_021913.5 | *JAK2* | NM_004972.4 | *BRCA1* | NM_007294.4 |
| *FANCC* | NM_000136.2 | *B2M* | NM_004048.4 | *KDR* | NM_002253.4 | *BRCA2* | NM_000059.3 |
| *FANCD2* | NM_033084.4 | *BIRC3* | NM_001165.5 | *KIT* | NM_000222.3 | *CBL* | NM_005188.3 |
| *FANCE* | NM_021922.2 | *BRAF* | NM_001374258.1 | *KMT2A* | NM_001197104.2 | *CDH1* | NM_004360.4 |
| *FANCF* | NM_022725.3 | *CALR* | NM_004343.4 | *KRAS* | NM_004985.5 | *CDK4* | NM_000075.3 |
| *FANCG* | NM_004629.1 | *CBLB* | NM_170662.5 | *MAP2K1* | NM_002755.4 | *CKDN2A* | NM_000077.4 |
| *FANCI* | NM_001113378.1 | *CCND1* | NM_053056.3 | *MAPK1* | NM_002745.5 | *CEBPA* | NM_004364.4 |
| *FANCL* | NM_018062.3 | *CCND2* | NM_001759.4 | *MDM2* | NM_002392.6 | *CHEK1* | NM_001114121.2 |
| *FANCM* | NM_020937.3 | *CCND3* | NM_001760.5 | *MET* | NM_000245.4 | *CHEK2* | NM_007194.4 |
| *LIG4* | NM_002312.3 | *CDK6* | NM_001145306.2 | *MLH3* | NM_001040108.2 | *DDX41* | NM_016222.3 |
| *HAX1* | NM_002382.4 | *CSF1R* | NM_001288705.3 | *MPL* | NM_005373.3 | *ETV6* | NM_001987.5 |
| *MEN1* | NM_130799.2 | *CSF3R* | NM_000760.4 | *MTOR* | NM_004958.4 | *FANCA* | NM_000135.3 |
| *MUTYH* | NM_001128425.1 | *CTCF* | NM_006565.4 | *MYC* | NM_002467.6 | *FH* | NM_000143.3 |
| *NPAT* | NM_002519.2 | *CTNNA1* | NM_001903.5 | *MYCN* | NM_005378.6 | *GATA2* | NM_032638.4 |
| *PAX5* | NM_016734.2 | *CTNNB1* | NM_001904.4 | *MYD88* | NM_002468.5 | *IKZF1* | NM_006060.6 |
| *PMS1* | NM_000534.4 | *CUX1* | NM_181552.4 | *NFE2L2* | NM_006164.5 | *MLH1* | NM_000249.3 |
| *PMS2* | NM_000535.6 | *CXCR4* | NM_003467.3 | *NOTCH1* | NM_017617.5 | *MRE11A* | NM_005591.3 |
| *POLD1* | NM_002691.3 | *DAXX* | NM_001141969.2 | *NOTCH2* | NM_024408.4 | *MSH2* | NM_000251.3 |
| *RAD50* | NM_005732.3 | *DDR2* | NM_006182.4 | *NPM1* | NM_002520.7 | *MSH6* | NM_000179.3 |
| *RTEL1* | NM_001283009.2 | *DICER1* | NM_177438.3 | *NRAS* | NM_002524.5 | *NBN* | NM_002485.4 |
| | NM_032957.5 | *DNMT3A* | NM_022552.5 | *PBRM1* | - | *NF1* | NM_000267.3 |
| *SAMD9L* | NM_152703.4 | *EGFR* | NM_005228.5 | *PDGFRA* | NM_006206.6 | *NF2* | NM_000268.3 |
| *SLX4* | NM_032444.3 | *EP300* | NM_001429.4 | *PDGFRB* | NM_002609.4 | *PALB2* | NM_024675.3 |
| *SRP72* | NM_006947.3 | *EPHA3* | NM_005233.6 | *PIK3CA* | NM_006218.4 | *POLE* | NM_006231.3 |
| *TERC* | NR_001566.1 | *EPHA5* | NM_001281766.3 | *PIK3CB* | NM_006219.3 | *POT1* | NM_015450.3 |
| *TMEM127* | NM_017849.3 | *ERBB2* | NM_004448.4 | *PIK3R1* | NM_181523.3 | *PTEN* | NM_000314.7 |
| *WRN* | NM_000553.5 | *ERBB3* | NM_001982.4 | *PLCG2* | NM_002661.5 | *PTPN11* | NM_002834.4 |
| *XRCC2* | NM_005431.2 | *ERBB4* | NM_005235.3 | *PPP2R1A* | NM_014225.6 | *RAD51* | NM_002875.4 |
| *XRCC3* | NM_005432.4 | *ERCC3* | NM_000122.2 | *PTCH1* | NM_000264.5 | *RAD51C* | NM_058216.3 |
| | | *ESR1* | NM_018010.4 | *RAD21* | NM_006265.3 | *RAD51D* | NM_002878.3 |
| | | *EZH2* | NM_004456.5 | *RB1* | NM_000321.3 | *RET* | NM_020975.5 |
| | | *FAT3* | NM_001367949.2 | *SETBP1* | NM_015559.3 | *RUNX1* | NM_001754.4 |
| | | *FBXW7* | NM_001349798.2 | *SF3B1* | NM_006842.3 | *SAMD9* | NM_017654.3 |
| | | *FGFR1* | NM_023110.3 | *SMARCB1* | NM_003073.5 | *SDHA* | NM_004168.4 |
| | | *FGFR2* | NM_000141.5 | *SMC3* | NM_005445.4 | *SDHAF2* | NM_017841.2 |
| | | *FGFR3* | NM_000142.5 | *SMO* | NM_005631.5 | *SDHB* | NM_003000.2 |
| | | *FLT3* | NM_004119.3 | *SRSF2* | NM_001195427.2 | *SDHC* | NM_003001.3 |
| | | *FOXL2* | NM_023067.4 | *STAT3,* | NM_139276.3 | *SDHD* | NM_003002.4 |
| | | *GNA11* | NM_002067.5 | *STAT5B* | NM_012448.4 | *SMAD4* | NM_005359.5 |
| | | *GNAQ* | NM_002072.5 | *TET2* | NM_001146069.2 | *STK11* | NM_000455.4 |
| | | *GNAS* | NM_000516.7 | *TSC1* | NM_000368.5 | *TERT* | NM_198253.2 |
| | | *GRIN2A* | NM_001134407.3 | *TSC2* | NM_000548.5 | *TP53* | NM_000546.6 |
| | | *H3F3A* | NM_002107.7 | *U2AF1* | NM_006758.3 | *VHL* | NM_000551.3 |
| | | *HIST1H3B* | NM_003537.4 | | | *WT1* | NM_024426.5 |
