## Supplemental Figure 2 for "Prevalence of incidental germline variants detected via tumor-only mesothelioma genomic profiling"

**Supp Figure 2. Timeline of Multiple Cancer Diagnoses in Patients with Germline Mutations**

Figure available upon email request to the corresponding author.

Mesothelioma Diagnosis    Other Malignancy Diagnosis    Deceased
