## Supplemental Figure 3 for "Prevalence of incidental germline variants detected via tumor-only mesothelioma genomic profiling"

**Supplementary Figure 3. Immunohistochemical Characteristics**

| Immunohistochemistry Characteristics | Patients with Pathogenic or Likely Pathogenic Germline Genetic Variants n | [%] | Patients without Pathogenic or Likely Pathogenic Germline Genetic Variants n | [%] | p-Value |
|---|---|---|---|---|---|
| | 25 | 16% | 136 | 86% | |
| **Tumor BAP1 Status** | | | | | |
| Retained | 6 | 24% | 49 | 36% | 0.295 |
| Lost | 17 | 68% | 69 | 51% | 0.277 |
| No Data | 2 | 8% | 18 | 13% | |
| **Tumor PD-L1 Status** | | | | | |
| Positive | 14 | 56% | 61 | 45% | 0.380 |
| Negative | 9 | 36% | 49 | 36% | 0.717 |
| No Data | 2 | 8% | 26 | 19% | 0.799 |
| **% of PD-L1 Positive Tumor Cells** | | | | | |
| 0-5% | 7 | 50% | 21 | 34% | 0.450 |
| 6-20% | 2 | 14% | 9 | 15% | 0.971 |
| 21-40% | 2 | 14% | 11 | 18% | 0.891 |
| 41-60% | 2 | 14% | 16 | 26% | 0.711 |
| >61% | 1 | 7% | 4 | 7% | 1.000 |
| **PD-L1 Staining Intensity** | | | | | |
| Weak | 5 | 36% | 32 | 52% | 0.506 |
| Weak to Moderate | 3 | 21% | 10 | 16% | 0.84 |
| Moderate | 3 | 21% | 6 | 10% | 0.65 |
| Moderate to Strong | 0 | 0% | 6 | 10% | - |
| Strong | 0 | 0% | 2 | 3% | - |
| No Data | 3 | 21% | 5 | 8% | |

| | **Pathogenic or Likely Pathogenic** | | | | | | | |
|---|---|---|---|---|---|---|---|---|
| | *ATM* | | *BAP1* | | *CHEK2* | | OTHER | |
| Immunohistochemistry | n | % | n | % | n | % | n | % |
| Characteristics | 3 | 17% | 8 | 28% | 6 | 33% | 5 | 28% |
| **Tumor BAP1 Status** | | | | | | | | |
| Retained | 1 | 33% | 0 | 0% | 0 | 0% | 2 | 40% |
| Lost | 2 | 67% | 7 | 88% | 5 | 83% | 2 | 40% |
| No Data | 0 | 0%% | 1 | 13% | 1 | 17% | 1 | 20% |
| **Tumor PD-L1 Status** | | | | | | | | |
| Positive | 2 | 67% | 4 | 50% | 6 | 100% | 2 | 40% |
| Negative | 1 | 33% | 3 | 38% | 0 | 0% | 3 | 60% |
| No Data | 0 | 0 | 1 | 13% | 0 | 0% | 0 | 0% |
| **% of PD-L1 Postive Tumor Cells** | | | | | | | | |
| 0-5% | 0 | 0% | 2 | 50% | 3 | 50% | 2 | 100% |
| 6-20% | 0 | 0% | 2 | 50% | 0 | 0% | 0 | 0% |
| 21-40% | 1 | 50% | 0 | 0% | 1 | 17% | 0 | 0% |
| 41-60% | 1 | 50% | 0 | 0% | 1 | 17% | 0 | 0% |
| >61% | 0 | 0% | 0 | 0% | 1 | 17% | 0 | 0% |
| **PD-L1 Staining Intensity** | | | | | | | | |
| Weak | 0 | 0% | 1 | 33% | 2 | 40% | 1 | 50% |
| Weak to Moderate | 0 | 0% | 0 | 0% | 3 | 60% | 0 | 0% |
| Moderate | 2 | 0% | 1 | 33% | 0 | 0% | 0 | 0% |
| Moderate to Strong | 0 | 100% | 0 | 0% | 0 | 0% | 0 | 0% |
| Strong | 0 | 0% | 0 | 0% | 0 | 0% | 0 | 0% |
| No Data | 0 | 0% | 1 | 33% | 1 | 17% | 1 | 50% |
