## Supplemental Figure 4 for "Prevalence of incidental germline variants detected via tumor-only mesothelioma genomic profiling"

**Supplementary Figure 4:** P/LP Somatic Variants Detected On Tumor NGS

| Patient | Gene | Variant | Tumor VAF | Gender | Site of Disease | Histology |
|---|---|---|---|---|---|---|
| 1 | ATM | c.4909_1G>T, p.? | 50% | Female | Peritoneal | Epithelioid |
| | BAP1 | c.740del, p.V247Gfs*2 | 31% | | | |
| 2 | ATM | c.5932G>T, p.E1978* | 44% | Male | Pleural | Epithelioid |
| | BAP1 | c.266del, p.N89Tfs*9 | 16% | | | |
| 3 | ATM | c.6154G>A, p.Q2052K | 48% | Male | Pleural | Epithelioid |
| | ATRX | c.6201dup, p.P2068Tfs*5 | 32% | | | |
| | BAP1 | c.659+1G>T, p.? | 56% | | | |
| | NF2 | c.448-2A>C, p.? | 7% | | | |
| | TP53 | c.532dup, p.H178Pfs*3 | 49% | | | |
| 4 | ATR | c.1327A>T, p.R443* | 54% | Male | Pleural | Epithelioid |
| | PALB2 | c.2587-1G>T, p.? | 8% | | | |
| | TP53 | Loss - Equivocal | - | | | |
| 5 | BAP1 | c.1717del, p.L573Wfs*3 | 78% | Female | Peritoneal | Epithelioid |
| 6 | BAP1 | c.1717del, p.L573Wfs*3 | 43% | Female | Pleural | Epithelioid |
| 7 | BAP1 | c.1717del, p.L573Wfs*3 | 51% | Female | Peritoneal | Epithelioid |
| | BAP1 | c.1949_1956delinsC, p.L650Pfs*3 | 18% | | | |
| 8 | BAP1 | c.1330del ,p.T44Pfs*127 | 25% | Male | Bicavitary | Epithelioid |
| | BAP1 | c.778C>T,e p.Q260* | 52% | | | |
| | CTNNA1 | c.468dup, p.V157Cfs*14 | 22% | | | |
| | NF1 | c.1844del, p.K615Sfs*16 | 5% | | | |
| 9 | BAP1 | c.376_377del, p.? | 46% | Male | Peritoneal | Epithelioid |
| | BAP1 | c.437+2A>T, p.? | 23% | | | |
| | CSF1R | c.2276_2280del, p.L756Pfs*23 | 18% | | | |
| 10 | BAP1 | c.272dup, p.C91Wfs*35 | 88% | Male | Peritoneal | Epithelioid |
| | BAP1 | Loss | - | | | |
| 11 | BAP1 | c.68-2A>G, p.? | 24% | Female | Pleural | Epithelioid |
| | TP53 | c.681dup, p.D228* | 20% | | | |
| 12 | BAP1 | c.178C>T, p.R60* | 46% | Male | Peritoneal | Epithelioid |
| | DDX3X | c.968C>T, p.T323I | 52% | | | |
| 13 | EPHA5 | c.511dup, p.Y171Lfs*5 | 8% | Male | Peritoneal | Epithelioid |
| 14 | BAP1 | c.37+9_122+60del, p.? | 8% | Male | Pleural | Epithelioid |
| 15 | BAP1 | c.354_358del, p.F118Lfs*6 | 21% | Male | Pleural | Biphasic |
| | CHEK2 | c.277del, p.W93Gfs*17 | 49% | | | |
| | DDX3X | c.1056_1061del, p,M352_D354delins | 34% | | | |
| 16 | BAP1 | c.581-2del, p.? | 76% | Male | Pleural | Epithelioid |
| | DDX3X | c.1438dup, p.R480Kfs*38 | 83% | | | |
| | NF2 | c.114G>A, p.E38E | 11% | | | |
| 17 | BAP1 | c.1321C>T, p.Q441* | 9% | Male | Pleural | Epithelioid |
| | CHEK2 | c.1100del, p.T367Mfs*15 | 42% | | | |
| | DDX3X | c.976C>T, p.R326C | 20% | | | |
| | NF2 | c.115G>A, p.E38E | 11% | | | |
| 18 | CHEK2 | c.1100del, p.T367Mfs*15 | 45% | Female | Pleural | Biphasic |
| | MET | c.3082+2T>C, p.? | 24% | | | |
| | NF1 | c.7188del, p.Y2398Tfs*20 | 9% | | | |
| 19 | - | None | - | Female | Peritoneal | Epithelioid |
| 20 | BAP1 | c.256-1_259del, p.? | 4% | Female | Pleural | Epithelioid |
| | DDX41 | c.490C>T, p.R164W | 46% | | | |
| 21 | NF2 | c.1009C>T, p.Q337* | 14% | Male | Pleural | Epithelioid |
| | TP53 | Loss - Equivocal | - | | | |
| 22 | BAP1 | c.2526-1G>A, p.? | 55% | Female | Pleural | Epithelioid |
| | CDKN2A | Loss - Equivocal | - | | | |
| | MRE11A | c.1222dup, p.T408Nfs*49 | 46% | | | |
| | NF2 | c.702_732del, p.G235Tfs*6 | 46% | | | |
| | TP53 | c.528C>G, p.C176W | 49% | | | |
| 23 | MSH6 | c.3261dup, p.F1088Lfs*5 | 22% | Male | Pleural | Epithelioid |
| | PTEN | c.493G>C, p.G165R | 49% | | | |
| 24 | BAP1 | Rearrangement | - | Male | Pleural | Epithelioid |
| 25 | CDKN2A | Rearrangement | - | Male | Peritoneal | Epithelioid |
| | NF1 | c.4947del, p.P1650Lfs*48 | 47% | | | |
| 26 | - | None | - | Female | Pleural | Epithelioid |
| 27 | BAP1 | Loss | - | Male | Pleural | Epithelioid |
| 28 | BAP1 | c.582del, p.W196Gfs*35 | 40% | Male | Pleural | Epithelioid |
| 29 | MLH3 | c.3367C>T, p.Q1123* | 49% | Male | Pleural | Epithelioid |
| | PTEN | c.121del, p.R41Dfs*13 | 26% | | | |
| | TP53 | c.818G>T, p.R273L | 43% | | | |
| 30 | - | None | - | Male | Peritoneal | Epithelioid |
| 31 | BAP1 | Loss/Rearrangement | - | Male | Pleural | Epithelioid |
| | TP53 | Loss | - | | | |
| 32 | BAP1 | c.253C>A< p.Q85K | 47% | Male | Pleural | Epithelioid |
| | DDX3X | c.546del, p.F182Lfs*39 | 54% | | | |
| 33 | BAP1 | c.2012_2013insAA, p.Tyr671* | 19% | Male | Pleural | Epithelioid |
| | NF2 | c.616G>T, p.E206* | 16% | | | |
| 34 | CDKN2A | Loss | - | Male | Pleural | Epithelioid |
| 35 | BAP1 | c.376-25_376del, p.? | 22% | Male | Peritoneal | Epithelioid |
| | BAP1 | c.583_591del, p.P195_G197del | 26% | | | |
| 36 | BAP1 | c.1909_1910del, p.K637Vfs*5 | 21% | Male | Pleural | Epithelioid |
| 37 | BRCA2 | c.8331+2T>C, p.? | 22% | Female | Pleural | Biphasic |
| 38 | - | None | - | Female | Peritoneal | Epithelioid |
| 39 | - | None | - | Male | Pleural | Epithelioid |
| 40 | BAP1 | 575del, p.D192Afs*39 | 14% | Male | Peritoneal | Epithelioid |
| 41 | DDX3X | c.1676T>A, p.L559H | 32% | Male | Peritoneal | Epithelioid |
| 42 | BAP1 | c.860C>G, p.S287* | 16% | Male | Pleural | Epithelioid |
| 43 | BAP1 | c.200A>G, p.D67G | 17% | Male | Peritoneal | Epithelioid |
| | DDX3X | c.1423C>T, p.R475C | 16% | | | |
| 44 | TP53 | Rearrangement | - | Female | Pleural | Epithelioid |
| 45 | BAP1 | Loss - Equivocal | - | Male | Pleural | Epithelioid |
| | BAP1 | Rearrangement | - | | | |
| | CDKN2A | Loss | - | | | |
| | NF2 | Loss | - | | | |
| 46 | BAP1 | Rearrangement | - | Male | TVT | Epithelioid |
| 47 | B2M | c.246_247del, p.F82Lfs*7 | 36% | Male | Pleural | Epithelioid |
| | CDKN2A | c.341C>T, p.P114L | 24% | | | |
| | NF2 | c.1246C>T, p.R416* | 34% | | | |
| | NRAS | c.182A>T, p.Q61L | 29% | | | |
| | TERT | c.-124C>T, p.= | 16% | | | |
| | TP53 | c.396G>C, p.K132N | 45% | | | |
| 48 | NF2 | c.105_106insT, p.N36* | 38% | Male | Peritoneal | Epithelioid |
| 49 | ARID2 | c.4732C>T, p.Q1578* | 43% | Male | Pleural | Epithelioid |
| | BAP1 | Rearrangement | - | | | |
| | CDKN2A | Loss | - | | | |
| | CUX1 | c.3327dup, p.D11110Rfs*65 | 9% | | | |
| 50 | TP53 | c.524G>A, p.R175H | 11% | Male | Pleural | Epithelioid |
| | TP53 | c.818G>A, p.R273H | 9% | | | |
| 51 | FBXW7 | c.2065C>T, p.R689W | 16% | Male | Pleural | Epithelioid |
| 52 | BAP1 | c.1679dup, p.L561Pfs*6 | 66% | Female | Pleural | Biphasic |
| | CDKN2A | Loss | - | | | |
| 53 | DDX3X | c.1102_1106delinsa, p.D368Ifs*11 | 23% | Male | Peritoneal | Epithelioid |
| 54 | NF2 | Loss - Equivocal | - | Male | TVT | Epithelioid |
| | STK11 | Rearrangement | - | | | |
| 55 | BAP1 | c.1360G>T, p.E454* | 41% | Male | Pleural | Epithelioid |
| | NF2 | c.276_299del, p.T93_F100del | 44% | | | |
| 56 | BAP1 | Loss | - | Male | Peritoneal | Epithelioid |
| | TP53 | c.155_158dup, p.W53* | 10% | | | |
| | TP53 | Loss - Equivocal | - | | | |
| 57 | CDKN2A | c.332del, p.G111Afs*35 | 20% | Male | Pleural | Sarcomtoid |
| | TERT | c.-124C>T, p.= | 22% | | | |
| | TP53 | c.734G>C, p.G245V | 26% | | | |
| 58 | BAP1 | Loss - Equivocal | - | Female | BIcavitary | Epithelioid |
| | STK11 | c.375-1C>T, p.? | 28% | | | |
| 59 | BAP1 | Loss | - | Male | Pleural | Epithelioid |
| | CDKN2A | Loss - Equivocal | - | | | |
| | TP53 | Loss - Equivocal | - | | | |
| 60 | BAP1 | Rearrangement | - | Female | Peritoneal | Epithelioid |
| | WT1 | c.1413_1432_55delinsT, p.? | 29% | | | |
| 61 | BAP1 | c.2057-2A>T, p.? | 40% | Male | Peritoneal | Epithelioid |
| 62 | - | None | - | Male | Pleural | Epithelioid |
| 63 | BAP1 | Loss | - | Female | Pleural | Epithelioid |
| 64 | BAP1 | c.178C>T, p.R60* | 21% | Male | Pleural | Epithelioid |
| | NF2 | c.949G>T, p.E317* | 22% | | | |
| | TP53 | Loss - Equivocal | - | | | |
| 65 | BAP1 | c.376-1G>A, p.? | 22% | Female | Pleural | Epithelioid |
| | NF2 | c.737del, p.P246Lfs*5 | 21% | | | |
| 66 | BAP1 | Loss | - | Male | Peritoneal | Epithelioid |
| | PBRM1 | c.899+2T>C, p.? | 52% | | | |
| 67 | BAP1 | c.2057073_2079del, p.? | 18% | Male | Pleural | Epithelioid |
| | TP53 | Loss - Equivocal | - | | | |
| 68 | BAP1 | c.579_580dup, p.G194Vfs*38 | 28% | Male | Peritoneal | Epithelioid |
| 69 | BAP1 | Loss - Equivocal | - | Female | Pleural | Epithelioid |
| | PBRM1 | Rearrangement | - | | | |
| 70 | BAP1 | c.1574_1599del, p.S525* | 86% | Male | Pleural | Biphasic |
| | CDKN2A | Rearrangement/large-scale deletion | - | | | |
| | PTEN | c.388C>G, p.R130G | 45% | | | |
| | PTEN | c.703dup, p.E235Gfs*8 | 9% | | | |
| 71 | CDKN2A | Loss - Equivocal | - | Female | Peritoneal | Epithelioid |
| | NF2 | c.114+1G>A, p.? | 11% | | | |
| 72 | BAP1 | c.580+2T>C, p.? | 20% | Female | Pleural | Sarcomtoid |
| | CDKN2A | Loss - Equivocal | - | | | |
| 73 | ATM | Rearrangement | - | Female | Peritoneal | Benign Multicystic |
| 74 | TERT | c.-124C>T, p.= | 16% | Female | Pleural | Epithelioid |
| | TP53 | Loss - Equivocal | - | | | |
| 75 | CDKN2A | c.173delinsCA, p.R58Pf*62 | 46% | Male | Pleural | Epithelioid |
| | KRAS | c.34G>T, p.G12C | 55% | | | |
| | TP53 | c.993+1G>A, p.? | 45% | | | |
| 76 | CHEK2 | c.920del, p.G307Efs*13 | 6% | Male | Pleural | Epithelioid |
| | NF2 | Rearrangement/large-scale deletion | - | | | |
| 77 | NF2 | c.51del, p.K17Nfs*8 | 22% | Male | Pleural | Biphasic |
| | TP53 | c.403T>C, p.C135R | 22% | | | |
| 78 | - | None | - | Female | Peritoneal | Epithelioid |
| 79 | BAP1 | c.666del, p.Y223Tfs*8 | 22% | Male | Peritoneal | Epithelioid |
| | NF2 | Rearrangement | - | | | |
| 80 | FAT3 | c.9193delinsTAT, p.R3065Yfs*15 | 6% | Male | Pleural | Epithelioid |
| | TP53 | Loss - Equivocal | - | | | |
| 81 | CDKN2A | Loss - Equivocal | - | Male | Pleural | Epithelioid |
| | NF2 | c.970C>T, p.Q324* | 8% | | | |
| | TERT | c.-146C>T, p.= | 16% | | | |
| 82 | TERT | c.-124C>T, p.= | 6% | Female | Pleural | Sarcomtoid |
| 83 | NF2 | c.855dup, p.N286* | 40% | Male | Pleural | Epithelioid |
| 84 | BAP1 | Loss | - | Male | Pleural | Epithelioid |
| | BAP1 | Rearrangement | - | | | |
| | CDKN2A | Loss | - | | | |
| 85 | FBXW7 | c.1394G>A, p.R465H | 25% | Female | Pleural | Epithelioid |
| | NF2 | c.1340+1G>T, p.? | 17% | | | |
| | PBRM1 | Rearrangement | - | | | |
| 86 | ARID2 | c.3565C>T, p.Q1189* | 15% | Male | Pleural | Sarcomtoid |
| | HNF1A | c.814C>T, p.R272C | 15% | | | |
| | MSH6 | c.3725G>A, p.R1242H | 14% | | | |
| | NF2 | c.193C>T, p.Q65* | 18% | | | |
| 87 | - | None | - | Male | Peritoneal | Epithelioid |
| 88 | BAP1 | c.438-1_452del, p? | 6% | Male | Peritoneal | Epithelioid |
| | BAP1 | c.86T>G, p.V29G | 50% | | | |
| 89 | - | None | - | Female | Peritoneal | Epithelioid |
| 90 | BAP1 | c.68-1G>C, p.? | 22% | Male | Pleural | Epithelioid |
| | H3F3A | c.110A>G, p.K37R | 18% | | | |
| 91 | BAP1 | Loss | - | Male | Peritoneal | Epithelioid |
| | CDKN2A | Loss - Equivocal | - | | | |
| | CDKN2A | Rearrangement/large-scale deletion | - | | | |
| | STAG2 | c.894-11_921del, p,? | 37% | | | |
| | TP53 | Loss - Equivocal | - | | | |
| 92 | BAP1 | c.1828A>T, p.R610* | 44% | Female | Pleural | Epithelioid |
| 93 | BAP1 | Rearrangement | - | Female | Pleural | Epithelioid |
| | NF2 | c.449_468del, p.T150Cfs*46 | 23% | | | |
| 94 | - | None | - | Male | Peritoneal | Epithelioid |
| 95 | - | None | - | Female | Pleural | Epithelioid |
| 96 | BAP1 | c.255G>T, p.Q85H | 18% | Female | Pleural | BIphasic |
| | TP53 | c.375G>A, p.T125T | 23% | | | |
| 97 | BAP1 | c.1063C>T, p.Q355* | 20% | Male | Pleural | Epithelioid |
| | CDKN2A | Rearrangement | - | | | |
| 98 | BAP1 | Reaarrangement | - | Female | Pleural | Epithelioid |
| | NF2 | c.1188del, p.K396fs*30 | 19% | | | |
| 99 | NF2 | c.364-2A>C, p.? | 24% | Male | Peritoneal | Epithelioid |
| 100 | BAP1 | c.1729+1G>A, p.? | 60% | Female | Bicavitary | Epithelioid |
| 101 | CDKN2A | Loss | - | Male | Pleural | Epithelioid |
| | NF2 | c.834_867del, p.K278Nfs*7 | 52% | | | |
| | NF2 | Loss - Equivocal | - | | | |
| | TP53 | Loss - Equivocal | - | | | |
| 102 | BAP1 | c.71_87delinsACA, p.V24Dfs*40 | 21% | Male | Pleural | Epithelioid |
| 103 | BAP1 | c.200A>G, p.D67G | 21% | Male | Pleural | Epithelioid |
| 104 | BAP1 | Loss | - | Female | Peritoneal | Epithelioid |
| | PBRM1 | Loss | - | | | |
| 105 | BAP1 | c.1941_1980delinsT, p.E647_F660delinsD | 54% | Male | Pleural | Epithelioid |
| | NF2 | c.655G>A, p. V219M | 37% | | | |
| | TP53 | c.154C>T, p.Q52* | 37% | | | |
| 106 | BAP1 | Loss | - | Female | Pleural | Biphasic |
| | BAP1 | Rearrangement/large-scale deletion | - | | | |
| | CDKN2A | Loss | - | | | |
| | TP53 | Loss - Equivocal | - | | | |
| 107 | TP53 | c.578A>G, p.H193R | 25% | Male | Pleural | Epithelioid |
| 108 | TP53 | Loss - Equivocal | - | Male | Pleural | Epithelioid |
| 109 | BAP1 | Loss | - | Female | Peritoneal | Epithelioid |
| | PBRM1 | Loss | - | | | |
| 110 | CDKN2A | Loss | - | Male | Pleural | Epithelioid |
| | TP53 | c.776A>T, p.D259V | 56% | | | |
| 111 | BAP1 | c.991_994del, .K331Pfs*3 | 38% | Male | Pleural | Epithelioid |
| | CDKN2A | Loss | - | | | |
| 112 | BAP1 | c.1152_1198del, p.S384Rfs*3 | 10% | Male | Peritoneal | Epithelioid |
| 113 | BAP1 | c.1638C>G, p.Y546* | 8% | Male | Pleural | Epithelioid |
| | BAP1 | c.203A>G, p.D68G | 8% | | | |
| | CDKN2A | Rearrangement | - | | | |
| 114 | TP53 | c.736A>G, p.M246V | 22% | Male | Pleural | Epithelioid |
| 115 | BAP1 | c.2186_2189delinsC, p.*729Qdelins*205 | 68% | Male | Pleural | Epithelioid |
| | CDKN2A | Loss | - | | | |
| | MDM2 | Amplification | - | | | |
| 116 | - | None | - | Male | Pleural | Epithelioid |
| 117 | TERT | c.-124C>T, p.= | 27% | Female | Pleural | Epithelioid |
| 118 | CDKN2A | Loss | - | Male | Pleural | Biphasic |
| 119 | - | None | - | Female | Pleural | Biphasic |
| 120 | CDKN2A | Rearrangement/Loss - Equivocal | - | Male | Pleural | Sarcomtoid |
| | NF2 | c.970del, p.Q324Rfs*22 | 48% | | | |
| | TERT | c.-124C>T, p.= | 55% | | | |
| 121 | BAP1 | c.1911_1930delinsTTCCTGC, p.K637N*14 | 74% | Male | Peritoneal | Epithelioid |
| | TP53 | Loss - Equivocal | - | | | |
| 122 | PIK3CA | c.1624G>A, p.E542K | 26% | Female | Pleural | Epithelioid |
| 123 | TP53 | c.637C>T, p.R213* | 37% | Male | Pleural | Epithelioid |
| 124 | CDKN2A | Loss - Equivocal | - | Male | Pleural | Sarcomtoid |
| | NF2 | c.1396C>T, p.R466* | 39% | | | |
| | TERT | c.-124C>T, p.= | 24% | | | |
| | TP53 | Loss | - | | | |
| 125 | - | None | - | Male | Peritoneal | Epithelioid |
| 126 | BAP1 | c.687C>G, p.N229K | 27% | Male | Pleural | Epithelioid |
| 127 | CDKN2A | Loss | - | Male | Pleural | Epithelioid |
| | TERT | Amplification - Equivocal | - | | | |
| 128 | BAP1 | Loss | - | Female | Bicavitary | Epithelioid |
| | MLH3 | c.3563C>G, p.S1188* | 54% | | | |
| 129 | CDKN2A | Loss | - | Male | Pleural | BIphasic |
| 130 | BAP1 | c.1177C>T, p.Q393* | 33% | Male | Pleural | Epithelioid |
| | CDKN2A | Loss | - | | | |
| | NF2 | Loss | - | | | |
| 131 | ASXL1 | c.1772dup, p.T591* | 6% | Male | Pleural | Epithelioid |
| | BAP1 | Loss - Equivocal | - | | | |
| | CDKN2A | Loss | - | | | |
| | NF2 | Rearrangement | - | | | |
| | NF2 | Loss | - | | | |
| 132 | - | None | - | Male | Pleural | Biphasic |
| 133 | BAP1 | c.1309_1311delinsT, p.L437Ffs*13 | 40% | Male | Pleural | Epithelioid |
| | CDKN2A | Loss | - | | | |
| | NF2 | c.784C>T, p.R262* | 66% | | | |
| 134 | NF2 | c.1446+1G>A, p.? | 24% | Female | Peritoneal | Epithelioid |
| 135 | KDM6A | Rearrangement | - | Female | Pleural | Epithelioid |
| | WT1 | Rearrangement | - | | | |
| 136 | BAP1 | Rearrangement | - | Female | Pleural | Biphasic |
| | MLH1 | Rearrangement | - | | | |
| | TP53 | c.380C>T, p.S127F | 42% | | | |
| 137 | - | None | - | Female | Pleural | Epithelioid |
| 138 | CDKN2A | Loss | - | Female | Peritoneal | Epithelioid |
| | NF2 | c.100dup, p.E34Gfs*15 | 29% | | | |
| 139 | - | None | - | Female | Peritoneal | Epithelioid |
| 140 | - | None | - | Male | Pleural | Epithelioid |
| 141 | - | None | - | Male | Pleural | Epithelioid |
| 142 | - | None | - | Male | Pleural | Epithelioid |
| 143 | CDKN2A | Loss | - | Male | Pleural | Epithelioid |
| | CS3FR | c.2259_2313del, p.R754Afs*8 | 21% | | | |
| | FBXW7 | c.585-60_605del, p.? | 17% | | | |
| | TP53 | Loss - Equivocal | - | | | |
| 144 | CDKN2A | Loss | - | Female | Pleural | Biphasic |
| | NF2 | c.241del, p.V81Yfs*42 | 55% | | | |
| | TP53 | c.666del, p.Y223Tfs*8 | 54% | | | |
| 145 | - | None | - | Female | Pleural | Epithelioid |
| 146 | PBRM1 | Loss | - | Female | Peritoneal | Epithelioid |
| 147 | - | None | - | Male | Pleural | Sarcomtoid |
| 148 | ARID2 | c.430C>T, p.Q144* | 38% | Female | Pleural | Epithelioid |
| | TERT | c.-146C>T, p.= | 24% | | | |
| | TP53 | c.158G>A, p.W53* | 14% | | | |
| | TP53 | c.524G>A, p.R175H | 6% | | | |
| | TP53 | c.536A>G, p.H179R | 12% | | | |
| 149 | TP53 | c.1025G>C, p.R342P | 8% | Male | Pleural | Epithelioid |
| 150 | CDKN2A | Rearrangement | - | Female | Pleural | Sarcomtoid |
| 151 | ATM | c.2239del, p.Q747Rfs*6 | 16% | Male | Peritoneal | Epithelioid |
| | BAP1 | c.1730-1G>T, p.? | 38% | | | |
| | NF2 | c.389del, p.K130Rfs*44 | 33% | | | |
| 152 | BAP1 | Loss | - | Female | Pleural | Epithelioid |
| | TP53 | Loss | - | | | |
| 153 | BAP1 | c.581-95_595del, p.? | 25% | Male | Pleural | Epithelioid |
| 154 | CDKN2A | Loss | - | Female | Peritoneal | Epithelioid |
| | NF2 | c.1126_1135del, p.S377Qfs*46 | 57% | | | |
| 155 | CDKN2A | Loss - Equivocal | - | Female | Pleural | Epithelioid |
| | NF2 | Loss | - | | | |
| 156 | MSH6 | c.3G>A, p.? | 8% | Male | Peritoneal | Epithelioid |
| | TERT | c.-124C>T, p.= | 5% | | | |
| 157 | CDKN2A | Loss | - | Female | Peritoneal | Epithelioid |
| | TP53 | c.422G>A, p.C141Y | 59% | | | |
| | TP53 | Loss - Equivocal | - | | | |
| 158 | BAP1 | Rearrangement | - | Male | Pleural | Epithelioid |
| 159 | DDX3X | c.1061A>T, p.D345V | 29% | Male | Pleural | Epithelioid |
| 160 | - | None | - | Female | Bicavitary | Epithelioid |
| 161 | BAP1 | c.553G>C, p.G185R | 32% | Male | Pleural | Epithelioid |
| | CDKN2A | Loss - Equivocal | - | | | |
